# Advanced Echocardiographic Assessment in Transthyretin Amyloidosis: Early Phenotype Markers in Mutation Carriers

**DOI:** 10.1101/2024.08.27.24311875

**Authors:** Lorena Squassante Capeline, Frederico José Neves Mancuso, Gardênia Silva Lobo Oishi, Marly Uellendahl, Fernando Naylor, Acary Souza Bulle Oliveira, Valdir Ambrósio Moises

**Affiliations:** Universidade Federal de São Paulo - Escola Paulista de Medicina, Disciplina de Cardiologia, Setor de Ecocardiografia, São Paulo, São Paulo, Brazil; Universidade Federal de São Paulo - Escola Paulista de Medicina, Disciplina de Emergência e Medicina Baseada em Evidências, São Paulo, São Paulo, Brazil; Universidade Federal de São Paulo, - Escola Paulista de Medicina, São Paulo, Brazil. Doenças Neuromusculares, Disciplina de Neurologia Clínica, Departamento de Neurologia e Neurocirurgia.

**Keywords:** cardiac amyloidosis, echocardiography, speckle-tracking, strain, cardiomyopathies, hereditary ATTR amyloidosis

## Abstract

**Background:** Transthyretin cardiac amyloidosis (ATTR-CA) is a life-threatening heart condition due to mutations in the transthyretin (TTR) gene. This study aimed to investigate ventricular global longitudinal strain (GLS) and global circumferential strain (GCS) alongside traditional echocardiographic measures in TTR carries with preserved ejection fraction (EF).

**Methods:** Individuals with a family history of ATTR-CA, positive genetic testing for TTR, no clinical cardiac involvement and preserved EF were matched with healthy controls.

**Results:** Sixty-five patients were included: 31 TTR+ and 34 controls. No significant differences in chamber dimensions or EF were observed between groups. However, septal velocities were markedly lower in TTR+ than controls (9 [7;12] cm/s vs. 12 [10;13] cm/s, p=0.003). Additionally, both GCS and GLS were significantly reduced in the TTR+ group compared to controls (-21 ±4% vs. -28 ±4%, p<0.001 and -19 ±3% vs. -25 ±2%, p<0.001, respectively). Furthermore, right ventricular strain showed a similar trend, being lower in the TTR+ group compared to controls (-18 ±4% vs. -25 ±5%, p<0.005). Logistic regression analysis identified GLS (OR = 2.46; 95% CI 1.30-4.62) and E/e’ mean ratio (OR = 2.04; 95% CI 1.09-3.91) as independent predictors of positive genetic testing for TTR.

**Conclusion:** Asymptomatic individuals with a family history of ATTR-CA and positive genetic testing for TTR, despite preserved EF, exhibit lower GLS and GCS compared to controls. GLS and the E/e’ ratio were identified as independent predictors of positive genetic testing for TTR.

## Introduction

Hereditary transthyretin amyloidosis (ATTRv) represents a complex group of autosomal dominant disorders characterized by the extracellular deposition of amyloid fibrils derived from misfolded transthyretin (TTR). This condition is associated with over 130 different mutations in the TTR gene, resulting in significant variability in genotype-phenotype expression. Recent advancements in treatment underscore the critical need for a standardized approach to patient management, particularly focusing on accurate disease diagnosis and timely intervention strategies ^1–4^.

The timing and criteria for offering genetic testing to at-risk family members remain uncertain and require consideration of multiple factors including the specific mutation, penetrance, age of onset, and severity of disease in affected relatives. Early identification of pathogenic ATTRv enables proactive screening of asymptomatic gene carriers, facilitating prompt diagnosis and initiation of disease-modifying therapies aimed at improving clinical outcomes ^5^.

Cardiac amyloidosis (CA), a notable cardiac manifestation of ATTRv, is characterized by the infiltration of amyloid deposits within the myocardium, leading to restrictive cardiomyopathy and significant implications for prognosis and therapeutic decisions ^6–8^. The severity of CA is typically evaluated based on parameters such as ventricular wall thickness, systolic, and diastolic function ^8^. While diagnostic strategies for symptomatic CA are well-established, approaches for detecting subclinical conditions remain limited ^3^. Given the nonspecific clinical presentation, a high index of suspicion is crucial for accurate diagnosis, often relying on non-invasive imaging techniques including echocardiography, cardiovascular magnetic resonance, and cardiac scintigraphy ^6^. Moreover, with specific treatments available, timely diagnosis of cardiac amyloidosis is crucial than ever.

Contemporary echocardiographic methods, particularly strain and strain rate analysis, have emerged as effective screening tools for detecting early stages of CA. Two-dimensional speckle tracking echocardiography measures myocardial deformation and has shown promise in distinguishing CA from other cardiovascular conditions ^9^. Unlike traditional measures such as ejection fraction or shortening fraction, which may remain preserved until late disease stages, speckle tracking echocardiography offers sensitivity to detect subtle myocardial abnormalities early in the disease process ^9^.

The aim of this study was to evaluate echocardiographic and myocardial deformation alterations in carriers of the transthyretin variant with preserved ejection fraction and a positive family history for transthyretin amyloid cardiomyopathy. Additionally, the study seeks to identify clinical and echocardiographic variables that predict the genetic diagnosis of ATTRv in this population.

## Methods

This cross-sectional study recruited individuals with a known family history of ATTR-CA and positive genetic testing for TTR variant between January 2018 and December 2019 at a single tertiary center. Inclusion criteria comprised the absence of clinical cardiac involvement and preserved ejection fraction. A control group, matched for age and sex, was selected from healthy individuals without a family history of ATTR-CA.

None of the participants exhibited symptoms such as dyspnea, palpitations, syncope, arrhythmias, edema, or signs of cardiac disease upon physical examination. These individuals were defined as asymptomatic transthyretin variant carriers (TTR+) from a cardiovascular standpoint. All participants underwent transthoracic 2-dimensional echocardiography (TTE) confirming preserved ejection fraction at the time of enrollment.

Exclusion criteria included patients with a cardiac rhythm other than sinus rhythm or electrocardiographic evidence of bundle branch block, uncontrolled systemic arterial hypertension, primary valvular disease, congenital heart disease, history of myocardial revascularization surgery, known coronary artery disease, inadequate TTE images for strain analysis, or a history of previous diseases affecting other organs that could potentially influence cardiac function.

All patients provided written informed consent to participate in the study, which adhered to the principles of the Declaration of Helsinki and was approved by the Institutional Research Ethics Committee.

### Clinical evaluation

Cardiological evaluation was conducted concurrent with TTE examinations. A comprehensive physical examination was performed, followed by standard electrocardiography and TTE. Laboratory analyses were not conducted specifically for the study.

### Genetic analysis

Prior to study inclusion, all patients underwent genetic analysis as per institutional protocol, conducted only after obtaining patient consent. Genetic testing involved sequencing genes associated with neuropathies. Variant classification followed the guidelines of the American College of Medical Genetics^10^.

### Echocardiography

TTE examinations were conducted using a high-quality echocardiographic system (CX 50, Philips Medical Systems, Andover, MA, USA) equipped with a 1 to 5 MHz transducer (S5-1). Patients were in the left lateral decubitus position for the examination. Measurements were performed in accordance with current guidelines from the European Association of Cardiovascular Imaging and American Society of Echocardiography ^11^.

Left ventricular (LV) measurements included assessment of LV interventricular septum thickness (IVSd) and posterior wall thickness (PWd) at end-diastole, as well as internal diastolic and systolic diameters. LV mass, LV mass index, and relative wall thickness were calculated and applied to classify LV geometry according to established guidelines. Systolic function was evaluated using LV ejection fraction determined by the biplane Simpson’s method ^11^.

LV diastolic function was assessed using several parameters: mitral valve inflow early (E) and late (A) diastolic velocities, E/A ratio, and deceleration time. Additionally, tissue Doppler imaging measured early diastolic velocities of the medial and lateral mitral annulus (e′), averaged to calculate mean E/e′ in accordance with established guidelines. Left atrial (LA) volume index and maximum systolic velocity of tricuspid regurgitation, if available, were also utilized to analyze diastolic function ^12^.

LA volume was measured using the biplane area-length method and indexed to body surface area to calculate the LA volume index. Right ventricular (RV) size was assessed by diastolic diameter, and right atrial size by systolic volume, both from the apical 4-chamber view, following recommended protocols^11^.

To assess LV and RV systolic strain, all imaging acquisitions were digitally recorded over five consecutive cardiac cycles and stored in raw format. Analysis of recorded images was conducted by a single experienced echocardiographer blinded to subject conditions using speckle-tracking method estimated by a commercially available software validated for this purpose (QLAB Version 10.5; Philips Medical Systems, Andover, MA, USA) ^11^. The end-diastole was considered as the first frame when the mitral valve was closed, and end-systole was defined when the aortic valve was closed ^13^. The software traced speckles along the endocardial and epicardial borders throughout the cardiac cycle, and the region of interest was adjusted to encompass the entire myocardium ^14^.

LV longitudinal strain (LS) was assessed from the three apical long-axis views (4-, 2-, and 3-chamber views), while circumferential strain (CS) was evaluated from three short-axis views (base, mid, and apex). Global longitudinal strain (GLS) and global circumferential strain (GCS) values were derived by averaging LS and CS values, respectively, from all tracked LV segments ^14^. In cases where tracking was challenging or segments were excluded due to insufficient quality, GLS and GCS were recalculated using the remaining segments to ensure consistency and reliability across analyses ^14^.

RV longitudinal strain was assessed using the apical four-chamber view focused on the RV. Segmental strain values were obtained for distinct RV segments (free wall and septum) based on tracked myocardial segments. Global RV longitudinal strain (RVGLS) was derived by averaging segmental strain values ^15^.

Established LV strain-derived variables were calculated: septal peak LS apical to basal segment ratio (SAB) using 4-chamber view values; relative apical sparing of GLS ratio (RELAPS), the average apical segments peak LS divided by average basal and mid segments peak LS from all three apical views ^9,16,17^. Published cut-offs for detecting CA are >2.1 for SAB, >1 for RELAPS ^9,16,17^.

### Statistical Analysis

Data were analyzed using SPSS software (IBM Corporation, version 18, New Orchard Road Armonk, NY). Continuous variables were expressed as mean ± standard deviation or median [interquartile range] and compared using Student’s t-test or Mann-Whitney test, as appropriate. Categorical variables were compared using the chi-square test. Fisher’s exact test was applied to analyze categorical variables, and the likelihood ratio test was used to compare distribution fits.

Univariate analysis encompassed clinical and echocardiographic variables, with variables showing statistical significance (p-value < 0.10) included in the multiple logistic regression model. A stepwise variable selection procedure was employed to determine independent predictors of positive genetic study for TTR. Receiver operating characteristic curves (ROC) were constructed to assess predictive accuracy, with calculation of the area under the curve (AUC). Optimal cut-off values were determined using the Youden index derived from ROC curves. P-value <0.05 was considered statistically significant.

Intra and interobserver variabilities of GLS were assessed using the Bland-Altman method in a subset of 10 randomly selected patients, 60 days after initial assessment, with examiners blinded to previous results and each other’s assessments. The analysis included determination of bias and limits of agreement to ensure reliability and consistency of measurements.

## Results

Out of an initial enrollment of 36 individuals with TTR+, five were excluded from the study: one due to atrial fibrillation, one with unstable angina diagnosed during consultation, one with decompensated hypertension without specific therapy, and two who had undergone liver transplants. Therefore, a total of 31 patients were eligible and enrolled (Figure 1). Controls consisted of 34 asymptomatic individuals without neurological disease, and with normal laboratory and noninvasive cardiac test results. Comprehensive details of all 65 subjects included in the study are presented in Table 1.

**Figure 1.**
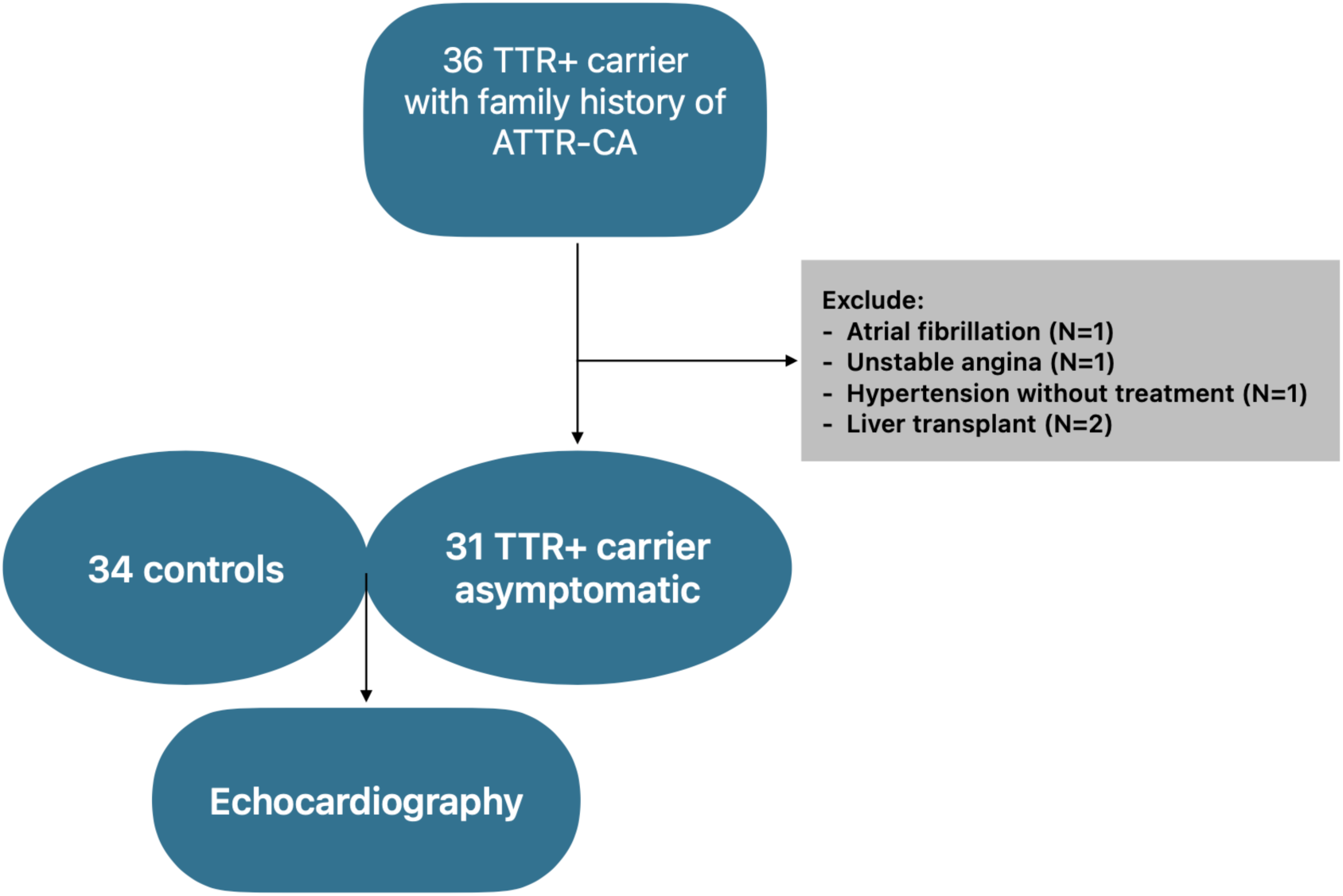
Population of the Study.

**Table 1.** Main Clinical Characteristics.

| Variable | Control (n = 34) | TTR+ (n = 31) | p-value |
| --- | --- | --- | --- |
| Male sex, n | 19 (55.9%) | 13 (41.9%) | 0.261<sup>a</sup> |
| Age, years | 37 (27; 41) | 40 (29; 57) | 0.090<sup>b</sup> |
| Weight, kg | 73 ± 13 | 74 ± 19 | 0.838<sup>c</sup> |
| Height, cm | 171 ± 8 | 169 ± 9 | 0.291<sup>c</sup> |
| BSA, m<sup>2</sup> | 1.8 ± 0.2 | 1.8 ± 0.2 | 0.609<sup>c</sup> |
| BMI, kg/m<sup>2</sup> | 25 ± 3 | 26 ± 5 | 0.423<sup>c</sup> |
| Obesity | 14 (41.2%) | 16 (51.6%) | 0.255<sup>d</sup> |
| Overweight | 11 (32.4%) | 9 (29.0%) |  |
| Obesity Grade I | 3 (8.8%) | 5 (16.1%) |  |
| Obesity Grade II | 0 (0%) | 2 (6.5%) |  |
| Systolic BP, mm Hg | 110 (100; 120) | 110 (110; 120) | 0.565<sup>b</sup> |
| Diastolic BP, mm Hg | 70 (60; 70) | 80 (60; 80) | 0.184<sup>b</sup> |
| Clinical history of hypertension | 3 (8.8%) | 2 (6.5%) | >0.999<sup>e</sup> |
| Symptoms | - | 14 (45.2%) | **<0.001<sup>a</sup>** |
| Polyneuropathy | - | 9 (29.0%) |  |
| Intestinal dysautonomia | - | 3 (9.6%) |  |
| Dysautonomia | - | 1 (3.2%) |  |
| Carpal tunnel syndrome | - | 1 (3.2%) |  |

P-values < 0.05 are show in bold. Variables are presented as mean ± SD, median (IQR), or percentile (%).
a: Chi-square, b: Mann-Whitney, c: Student's t-test, d: Fisher's exact, e: Likelihood ratio.
Abbreviation: BMI: Body Mass Index; BP: Blood pressure; BSA: Body Surface Area.

There were no significant differences between the groups in terms of sex, age, anthropometric measurements, obesity prevalence, blood pressure levels, or the prevalence of arterial hypertension. Among the 31 patients with TTR+, 14 (45.2%) exhibited extracardiac symptoms during their cardiological assessment. Specifically, polyneuropathy was observed in nine (29%) individuals, intestinal symptoms in three (9.6%), and carpal tunnel syndrome and dysautonomia in one (3.2%) individual each (Table 1).

### Genetic Characteristics

Among the 31 carriers of the TTR variant with preserved ejection fraction and a positive family history for ATTR-CA, the Val50Met variant was identified in 23 patients (74.1%). The Ile127Val variant was detected in four patients (13%), while the Glu109Lys variant was found in two patients (6.5%). Additionally, the Val142Ile and Ala117Ser variants each occurred in one case (3.2%).

### Echocardiographic parameters

The main echocardiographic findings are summarized in Table 2. All individuals with TTR+ had LV internal diameters within the normal range based on reference values. However, individuals with TTR+ exhibited significantly greater IVSd and PWd compared to controls (Table 2). Among the 31 TTR+ individuals, nine women had an IVSd greater than 9 mm, while five men had a IVSd greater than 10 mm; additionally, five women had a PWd greater than 9 mm, and four men had an PWd greater than 10 mm. LV mass, LV mass indexed, and LV relative wall thickness were significantly higher in the TTR+ group compared to controls; alterations in LV geometric pattern were also more frequent in the TTR+ group than in controls (p < 0.001) (Table 2). Among TTR+ individuals, LV concentric remodeling occurred in eight (four men and four women), LV concentric hypertrophy was observed in four (one man and three women), and LV eccentric hypertrophy in one individual. In the control group, only two individuals had LV concentric remodeling (Table 2).

**Table 2.**
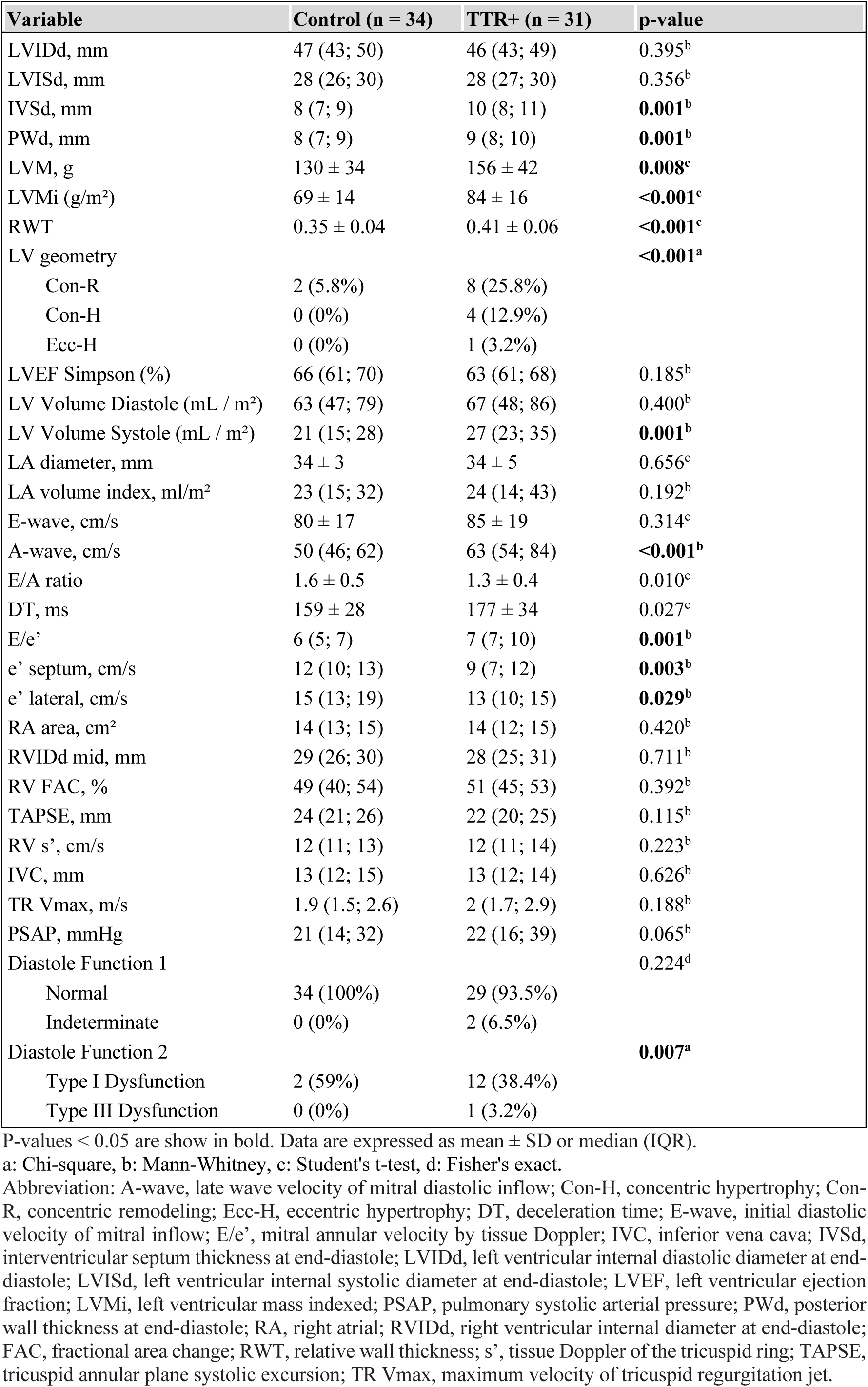
Echocardiographic Parameters.

LV ejection fraction did not differ significantly between the groups. Among the parameters assessed for LV diastolic function, the septal and lateral e’ waves and E/A ratio were significantly lower in TTR+ individuals compared to controls, although these values remained within the normal range (Table 2). The E/e’ ratio was significantly higher in the TTR+ group compared to controls.

Considering the majority of individuals had normal LV geometry, initial guideline-based analysis indicated normal diastolic function in the control group, with two individuals in the TTR+ group showing indeterminate diastolic function; however, this difference was not statistically significant. When considering only individuals with altered LV geometry, there were significantly more cases of altered diastolic function in the TTR+ group (13 out of 31 individuals) compared to controls (two out of 34 individuals) (p < 0.007). Among TTR+ individuals, 12 had type I diastolic dysfunction and one had type III. In the control group, both individuals exhibited type I dysfunction (Table 2).

### Myocardial deformation

GCS was markedly reduced in TTR+ individuals compared to controls, measuring -21 ± 4% versus -28 ± 4%, respectively (p < 0.001) (Table 3). In TTR+ group, four individuals exhibited GCS values below -18%, whereas in the control group, only one individual had a GCS of -18%.

**Table 3.** Myocardial Strain Parameters.

| Variable | Control (n = 34) | TTR+ (n = 31) | p-value |
| --- | --- | --- | --- |
| GCS*, % | -28 ± 4 | -21 ± 4 | **<0.001<sup>c</sup>** |
| CS basal*, % | -26 ± 5 | -18 ± 4 | **<0.001<sup>c</sup>** |
| CS medium*, % | -27 ± 5 | -20 ± 4 | **<0.001<sup>c</sup>** |
| CS apical*, % | -31 ± 6 | -24 ± 6 | **<0.001<sup>c</sup>** |
| GLS, % | -25 (-26; -23) | -19 (-20; -17) | **<0.001<sup>b</sup>** |
| LS basal, % | -22 ± 3 | -17 ± 3 | **<0.001<sup>c</sup>** |
| LS medium, % | -24 ± 3 | -18 ± 3 | **<0.001<sup>c</sup>** |
| LS apical, % | -27 ± 4 | -23 ± 4 | **<0.001<sup>c</sup>** |
| 4C, % | -24 (-26; -21) | -19 (-21; -17) | **<0.001<sup>b</sup>** |
| 2C, % | -24 (-26; -22) | -19 (-22; -17) | **<0.001<sup>b</sup>** |
| 3C, % | -24 (-27; -23) | -19 (-21; -17) | **<0.001<sup>b</sup>** |
| RV GLS, % | -25 ± 5 (n=11) | -18 ± 4 (n=11) | **0.005<sup>c</sup>** |
| RV FW GLS, % | -24 ± 4 (n=11) | -18 ± 4 (n=11) | **<0.001<sup>c</sup>** |
| RELAPS | 0.58 ± 0.08 | 0.66 ± 0.08 | **<0.001<sup>c</sup>** |
| SAB | 1.4 ± 0.3 | 1.7 ± 0.3 | **<0.001<sup>c</sup>** |

P-values < 0.05 are show in bold. Data are expressed as mean ± SD or median (IQR).
b: Mann-Whitney, c: Student's t-test.
Abbreviation: 4C: four-chambers; 2C: two-chambers; 3C: three-chambers; CS: circumferential strain; FW: free wall; GCS: global circumferential strain; GLS: global longitudinal strain.
\*One case was excluded from the analysis due to circumferential strain reading limitation.

GLS was also significantly lower in TTR+ individuals compared to controls, -19 ± 3% versus -25 ± 2%, respectively (p < 0.001). Specifically, eight cases in the TTR+ group and one in the control group had GLS values less than -18%; likewise, five cases in the TTR+ group had GLS values less than -16%.

Additionally, CS and LS values at basal, mid, and apical regions were notably diminished in the TTR+ group compared to the Control group (Figure 2). When comparing CS and LS values among basal, mid, and apical regions within the same group, significant differences were observed. In the TTR+ group, CS increased significantly from the basal region (-18 ± 4%) to the mid region (-20 ± 4%) with a p-value of 0.002. The apical region had the highest CS at -24 ± 6%, significantly different from both the basal and mid regions (p < 0.001). For LS, the apical region showed higher values at -23 ± 4% compared to the basal (-17 ± 3%) and mid (-18 ± 3%) regions, with significant differences between apical and both basal and mid (p < 0.001). In the Control group, CS increased from the basal region (-26 ± 5%) to the mid region (-27 ± 5%), but this difference was not significant (p = 0.094). The apical region had the highest CS at -31 ± 6%, significantly different from both the basal and mid regions (p < 0.001). For LS, values increased from basal (-22 ± 3%) to mid (-24 ± 3%), but this difference was not significant (p = 0.02). The apical LS was higher at -27 ± 4% compared to basal and mid regions, with significant differences (p < 0.001).

**Figure 2.**
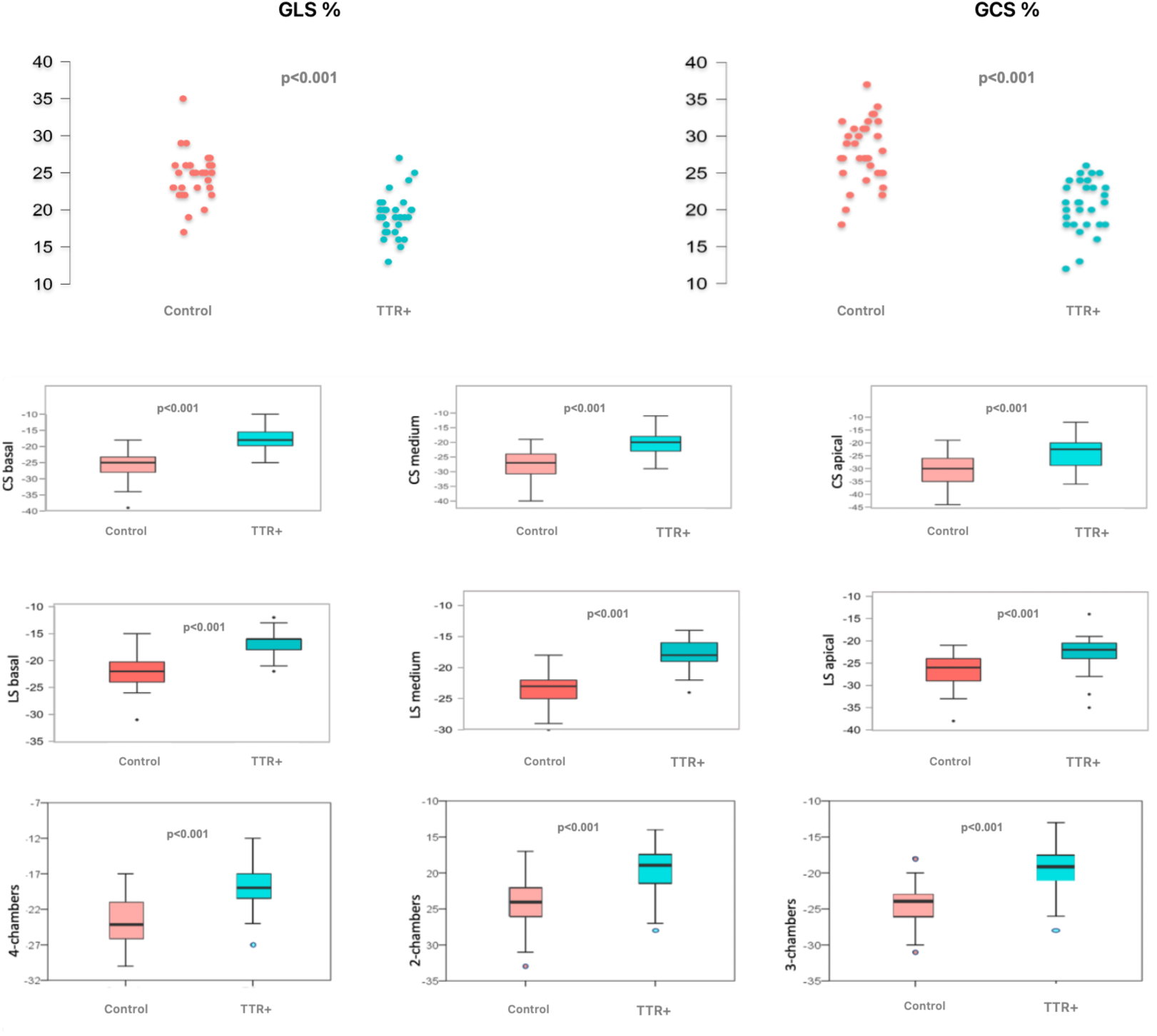
Strain values in TTR+ and control groups. LS, longitudinal strain; CS, circumferential strain; GLS. Global longitudinal strain; GCS, global circumferential strain.

RVGLS was assessed in eleven TTR+ individuals and eleven controls (Table 3). RVGLS was significantly lower in the TTR+ group compared to the control group (-18 ± 4% versus -25 ± 5%). A similar pattern was observed in the assessment of RV free wall GLS, with values of -18 ± 4% in the TTR+ group versus -24 ± 4% in the control group.

In the TTR+ group, RELAPS was significantly elevated at 0.66 ± 0.08 compared to 0.58 ± 0.08 in the control group (p < 0.001), and the SAB index was significantly elevated at 1.7 ± 0.3 compared to 1.4 ± 0.3 in the control group, with a p-value less than 0.001 (Table 3).

The comparison of TTR+ individuals with and without extracardiac symptoms no significant differences in GCS and GLS values were observed. Specifically, GCS was -21 ± 3% in asymptomatic individuals and -20 ± 3% in symptomatic individuals (p = 0.510). Similarly, GLS was -20 ± 3% in asymptomatic individuals and -19 ± 2% in symptomatic individuals (p = 0.322).

ROC analysis identified an optimal LV GLS cutoff of -21.5% to distinguish TTR+ individuals from controls, achieving a sensitivity of 87.1% and specificity of 91.2% (AUC = 0.890, 95% CI: 1.31 to 4.62, p < 0.005). Additionally, E/e’ ratio of 6.5 demonstrated a sensitivity of 80.6% and specificity of 64.7% for the same discrimination (AUC = 0.890, 95% CI: 1.31 to 4.62, p < 0.005) (Figure 3).

**Figure 3.**
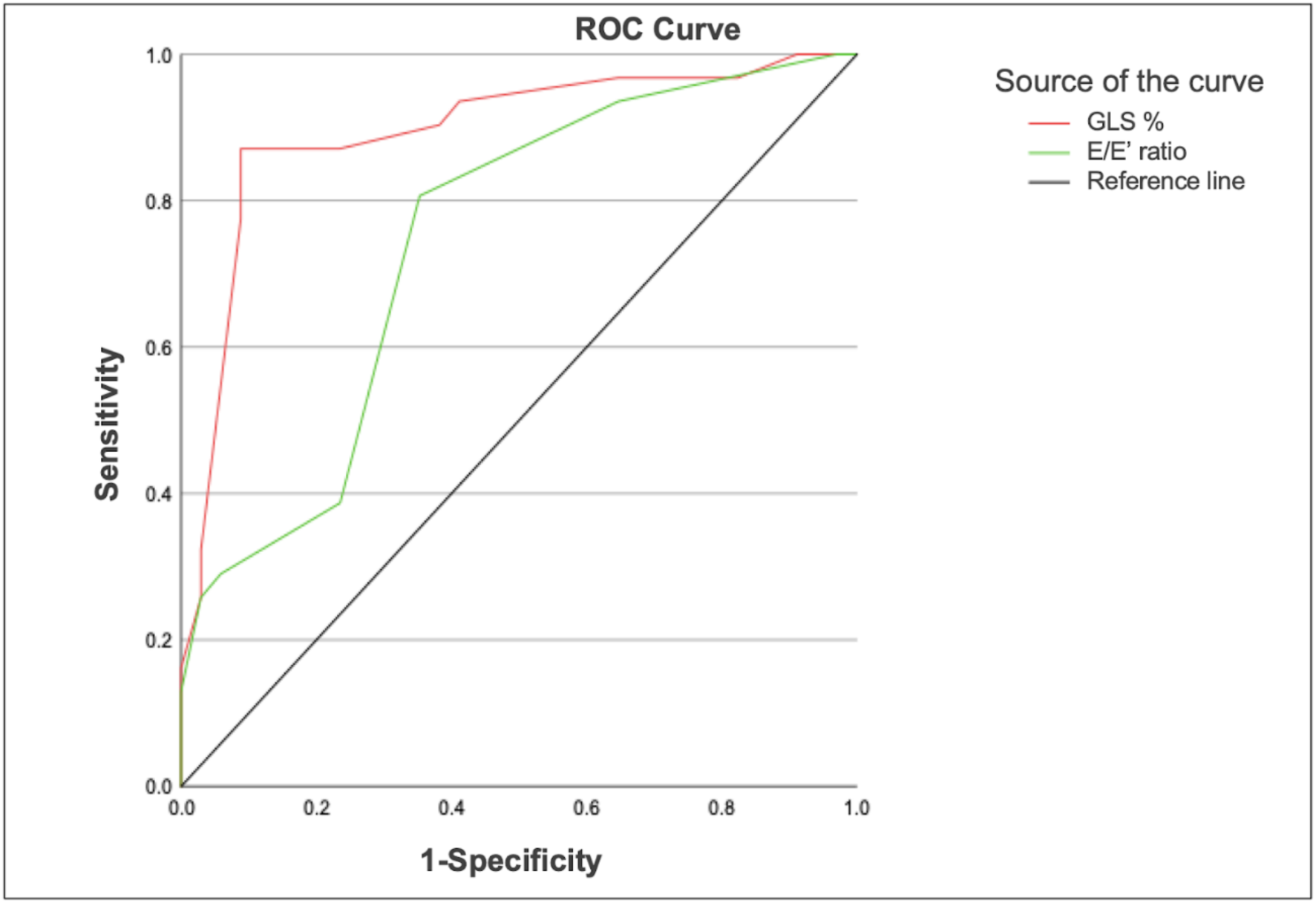
ROC analysis showed global longitudinal strain and E/e’ ratio to distinguish TTR+ carriers.

### Reproducibility

The study demonstrated strong correlations and high reproducibility with minimal bias. Interobserver variability had a bias of 0.235 and limits of agreement (±1.96 SD) from -3.97 to 4.17 (p = 0.882), while intra-observer variability had a bias of 0.278 with limits of agreement from -2.62 to 2.42 (p = 0.811).

## Discussion

This study provides compelling evidence that transthyretin variant carriers exhibit impaired myocardial deformation despite having preserved ejection fraction. Notably, our findings highlight a critical gap in the literature, as no previous study has specifically emphasized the characteristics of carriers in this context. Prior research has typically compared LV deformation in patients with ATTRv against those with conditions such as aortic stenosis, hypertrophic cardiomyopathy, or advanced-stage cardiac amyloidosis ^3,16,17^. Our study uniquely addresses this oversight by concentrating on transthyretin variant carriers, offering new insights into their myocardial deformation profile.

While myocardial biopsy remains the gold standard for definitively diagnosing ATTR-CA, distinguishing it from other causes of increased myocardial thickness in early stages presents ongoing clinical challenges ^1,18^. These findings emphasize the importance of heightened awareness and early detection strategies to improve outcomes for individuals with genetic predispositions to cardiac amyloidosis.

In this study, both groups showed comparable variables, except for the presence of extracardiac symptoms observed in some TTR+ individuals. The genetic variants identified in our study align with findings in the literature, indicating higher prevalence among American and Portuguese ethnicities ^19^. This underscores the diverse distribution of genetic variants associated with ATTR and highlights the importance of considering ethnic variability in genetic studies of this condition.

Among the 31 TTR+, 21 demonstrated alterations in myocardial thickness and LV geometric patterns despite the absence of cardiovascular symptoms. Specifically, five individuals exhibited LV hypertrophy, with four displaying predominantly concentric hypertrophy and one showing eccentric hypertrophy. Additionally, eight individuals showed minimal septal thickness increase, while another eight exhibited concentric remodeling. A previous study underscored increased LV mass and functional myocardial impairment in advanced ATTR-CA, even with preserved LV ejection fraction ^16^. Early stages may mimic hypertensive heart disease or other myocardial disorders. However, among TTR+ patients, only two had hypertension, suggesting early ATTR-CA manifestations, such as increased myocardial thickness and higher LV geometric alterations, may occur independently of hypertensive. Amyloid deposition in ATTR-CA often targets the interventricular septum, which may explain these findings ^9,16,20^.

All TTR+ in this study exhibited normal LV ejection fraction. According to diastolic function guidelines, assessing diastolic function in individuals with normal systolic function includes evaluating septal or lateral e’ waves, E/e’ ratio, LA volume index, and tricuspid regurgitation velocity ^12^. None of the participants met the criteria for diastolic dysfunction by this assessment, with diastolic function being indeterminate in two individuals. Nevertheless, given the potential for myocardial disease in these individuals, a secondary analysis commonly used in patients with heart disease or systolic dysfunction revealed a higher prevalence of diastolic dysfunction among TTR+ compared to controls: 12 individuals exhibited type I diastolic dysfunction, typically asymptomatic with normal LA pressure, while one displayed type III dysfunction, indicating elevated LA pressure. Previous research has shown that early-stage ATTR-CA involves myocardial amyloid deposition, leading to myocardial remodeling and fibrosis that can impair diastolic relaxation ^7^. In this study, this pattern was observed in 12 out of 31 TTR+. Diastolic filling parameters are known to carry prognostic significance in ATTR-CA, with increased amyloid infiltration potentially causing a restrictive pattern of LV filling ^2,7,17^.The data from this study suggest that reductions in strain may occur early in the disease among carriers of the ATTRv. These individuals exhibited significantly lower values of RV strain, LV GCS and GLS compared to controls. Similar findings were observed when comparing each LV regions between TTR+ and control. Rapezzi et al. have described increased myocardial thickness, altered diastolic function, and pericardial effusion as common cardiac changes in advanced stages of ATTR-CA, complicating treatment and potentially reducing survival ^8^.

In both study groups, LS and CS values progressively increased from the basal to the apical regions. However, values in each region were significantly lower in TTR+ compared to controls. RELAPS and SAB indices also differed between groups, suggesting potentially greater involvement of the mid and basal LV region in TTR+. This strain distribution in TTR+ individuals mirrors patterns described by Phelan et al. in advanced stages of ATTR-CA ^9^. These authors suggested that lower values of the basal and mid regions of the LV may result from more intense amyloid deposition in these areas. Importantly, Phelan et al. compared patients with ATTR-CA to those with hypertrophic cardiomyopathy and aortic stenosis, rather than to a control group without apparent myocardial disease, as in the present study ^9^. Cotella et al. in a recent publication including large number of patients highlight that the apical sparing does not have a high specificity for diagnosing cardiac amyloidosis ^21^. However, our study revealed that these TTR+ patients show early differences of myocardial deformation compared to controls with some regional left ventricular differences. This earlier stage of cardiac involvement underscores the importance of assessing not only the apical sparing pattern but also other echocardiographic variables, including parameters of the diastolic function of the left ventricle. These analyses can identify more subtle forms of myocardial changes in TTR+ carriers, facilitating early diagnosis and contributing to the prognosis of these individuals.

Furthermore, there were no significant differences in LV CS and LS values between TTR+ individuals with or without extracardiac symptoms, suggesting that neurological symptoms may not predict LV strain alterations.

RV strain, whether including the interventricular septum or not, behaved similarly to LV strain. In this study, four individuals in the TTR+ group had RV strain values below -16%, considered abnormal. Histopathological studies have shown a predilection for amyloid fibril deposition in the basal region of the interventricular septum ^22,23^. This raises questions about including or excluding the septum in RV strain analysis, with some arguing that including the septum could complicate RV strain analysis and advocating for analyzing only the RV free wall ^24^. In this study, RV GLS did not show significant difference regardless of whether the septum was included or not.

Several variables in this study were significantly linked to a TTR+ genetic test. However, logistic regression identified only GLS (OR = 2.46), and E/e’ ratio (OR = 2.07) as positively related to a TTR+ genetic test. GLS ≤ -21.5% distinguished TTR+ patients from the control group with a sensitivity of 87.1% and specificity of 91.2%. Similarly, E/e’ ratio ≥ 6.5 had a sensitivity of 80.6% and specificity of 64.7%. Chuy et al. demonstrated that GLS and E/e’ ratio are associated with overall survival in patients studied, providing incremental prognostic value in advanced stages of the disease ^25^.

The diagnostic challenge of ATTR-CA is compounded by the low penetrance of the genetic variant and subtle multi-organ involvement, equally amyloid fibril deposition progressively impacts tissues, with cardiac complications being a major cause of mortality ^26^. Therefore, early detection of cardiac involvement is critical for timely intervention and improved patient outcomes. The greatest clinical utility of our study is that it identified an early possible phenotypic diagnosis and may assist in the treatment effort to delay the clinical course of the disease.

## Limitations

There are some limitations to consider in this study. Firstly, it was a cross-sectional observational study conducted at a single tertiary referral center, involving a relatively small cohort of patients at various stages of polyneuropathy manifestation. However, given the low prevalence of confirmed cardiac amyloidosis in the general population, the number of participants in this study compares favorably with previously published research ^3,16,17^.

Furthermore, the strain analysis was conducted using equipment from a single supplier. Therefore, the findings may not be applicable to other commercially available strain systems across all vendors, although this limitation has not been found to be restrictive. Additionally, establishing a normative reference value for strain in clinical practice remains unclear and is dependent on specific clinical contexts ^27^.

Data collection and analysis were conducted by a single observer, which could potentially induce analysis bias. To minimize this possibility, images were recorded and saved without patient identification or group affiliation. Intra- and inter-observer analyses were performed at least 60 days after the initial image acquisition, and no significant differences were observed in both analyses.

While additional evaluation with cardiac magnetic resonance (CMR), scintigraphy, or laboratory exams could enhance identification of myocardial amyloid infiltration and diagnosis of cardiac amyloidosis, these measures were not included in the present study. Although CMR is diagnostically valuable, it carries risks such as allergic reactions to contrast agents, challenges for patients with claustrophobia, and its relatively high cost and invasive nature. Similarly, scintigraphy, though effective for detecting amyloid deposits, also involves exposure to radiation and may not be accessible in all clinical settings due to its cost and the need for specialized equipment.

It is notable that most patients in this study had echocardiographic values within normal limits, although significant differences compared to the control group were observed, and patients had no cardiovascular symptoms. Long-term follow-up with clinical and echocardiographic data could provide further insights into the implications of these findings over time.

## Conclusion

Our study demonstrated that the GLS and GCS of the left ventricle in carriers of the transthyretin variant with preserved ejection fraction and a positive family history for ATTR-CA were lower compared to the control group. Among the clinical and echocardiographic variables evaluated, GLS and the mean E/e’ ratio were independent predictors of positive genetic testing for transthyretin amyloidosis.

## Data Availability

All data produced in the present study are available upon reasonable request to the authors.

https://repositorio.unifesp.br/server/api/core/bitstreams/bd2ced50-5d5f-4ca6-b46a-52359f638216/content

